# Screening for Post-Traumatic Stress Disorder following Childbirth using the Peritraumatic Distress Inventory

**DOI:** 10.1101/2023.04.23.23288976

**Authors:** Kathleen M. Jagodnik, Tsachi Ein-Dor, Sabrina J. Chan, Adi Titelman Ashkenazy, Alon Bartal, Sharon Dekel

**Affiliations:** Department of Psychiatry, Harvard Medical School, Boston, Massachusetts; Department of Psychiatry, Massachusetts General Hospital, Boston, Massachusetts; School of Business Administration, Bar-Ilan University, Ramat Gan, Israel; School of Psychology, Reichman University, Herzliya, Israel

**Keywords:** Assessment, Delivery, Childbirth-related PTSD (CB-PTSD), Factorial Analysis, Maternal Mental Health, Mental Health Assessment, PTSD Following Childbirth (PTSD-FC), Postpartum, Birth Trauma, Maternal Morbidity

## Abstract

**Background:** Maternal psychiatric morbidities include a range of psychopathologies; one condition is post-traumatic stress disorder (PTSD) that develops following a traumatic childbirth experience and may undermine maternal and infant health. Although assessment for maternal mental health problems is integrated in routine perinatal care, screening for maternal childbirth-related PTSD (CB-PTSD) remains lacking. Acute emotional distress in response to a traumatic event strongly associates with PTSD. The brief 13-item Peritraumatic Distress Inventory (PDI) is a common tool to assess acute distress in non-postpartum individuals. How well the PDI specified to childbirth can classify women likely to endorse CB-PTSD is unknown.

**Objectives:** We sought to determine the utility of the PDI to detect CB-PTSD in the early postpartum period. This involved examining the psychometric properties of the PDI specified to childbirth, pertaining to its factorial structure, and establishing an optimal cutoff point for the classification of women with high vs. low likelihood of endorsing CB-PTSD.

**Study Design:** A sample of 3,039 eligible women who had recently given birth provided information about their mental health and childbirth experience. They completed the PDI regarding their recent childbirth event, and a PTSD symptom screen to determine CB-PTSD. We employed Exploratory Graph Analysis (EGA) and bootstrapping analysis to reveal the factorial structure of the PDI and the optimal PDI cutoff value for CB-PTSD classification.

**Results:** Factor analysis of the PDI shows two strongly correlated stable factors based on a modified 12-item version of the PDI consisting of (1) negative emotions and (2) bodily arousal and threat appraisal in regard to recent childbirth. This structure largely accords with prior studies of individuals who experienced acute distress resulting from other forms of trauma. We report that a score of 15 or higher on the modified PDI produces strong sensitivity and specificity. 88% of women with a positive CB-PTSD screen in the first postpartum months and 93% with a negative screen are identified as such using the established cutoff.

**Conclusions:** Our work reveals that a brief self-report screening concerning a woman’s immediate emotional reactions to childbirth that uses our modified PDI tool can detect women likely to endorse CB-PTSD in the early postpartum period. This form of maternal mental health assessment may serve as the initial step of managing symptoms to ultimately prevent chronic symptom manifestation. Future research is needed to examine the utility of employing the PDI as an assessment performed during maternity hospitalization stay in women following complicated deliveries to further guide recommendations to implement maternal mental health screening for women at high risk for developing CB-PTSD.

## Introduction

Each year, ∼4 million American women give birth. While most have healthy deliveries, 20%-30% undergo a highly stressful childbirth experience.^1,2^ A significant minority experience potentially life-threatening events and even maternal “near-miss” (nearly escaping death) in relation to childbirth, with rising rates in the U.S.^3^

Increasing evidence reveals that women can develop post-traumatic stress disorder in response to childbirth (CB-PTSD).^4^ Maternal CB-PTSD is estimated to occur in 3%-6% of the postpartum population^5,6^ and in nearly 20% of those who experience complicated deliveries.^7,8^ In the US, CB-PTSD affects ∼120,000-240,000 American women annually.^6^

CB-PTSD can become a debilitating condition of the postpartum period and consequently impair the health of the infant.^9^ A core feature of CB-PTSD is heightened physiological reactivity to childbirth reminders.^4^ This suggests that the child themselves can become a cue of the trauma and can trigger maternal distress, to his or her own detriment. Problems in the early formation of maternal-infant attachment are strongly associated with CB-PTSD,^1,10^ and interruption in attachment relations increases risk for behavior problems in children^11^ and mental illness in the adult offspring.^12,13^ CB-PTSD can further result in avoidance of future pregnancies.^14^

Currently, CB-PTSD is regarded an underdiagnosed and, consequently, undertreated maternal psychiatric morbidity.^15^ Because the symptoms of CB-PTSD are triggered in response to the childbirth event, the disorder has a clear onset. This offers a unique window of opportunities to identify postpartum women with traumatic stress reactions before a formal diagnosis can be confirmed, to increase the odds of potentially preventing CB-PTSD.

In non-postpartum samples, initial distress reactions to trauma exposure are well established to contribute to PTSD development,^16^ which is understood as a failure to extinguish acute stress responses.^17,18^ Peritraumatic (acute) subjective distress is an even stronger risk factor for PTSD than objective (stressor) elements of trauma.^19^ Similarly, reactions of acute distress in response to childbirth, e.g., fear and perceived loss of control, are associated with CB-PTSD.^1,6,20^ These reactions appear to strongly predict CB-PTSD more than obstetrical interventions and/or complications,^6,21^ and hence, constitute important information to identify women who are likely to develop CB-PTSD.

One of the most widely used measures to assess for acute stress reactions that may detect early signs of PTSD is the Peritraumatic Distress Inventory (PDI),^22^ a brief 13-item patient self-report questionnaire measuring emotional and physiological responses experienced during and shortly after a specified potentially traumatic event. The PDI has good psychometric properties in studies of individuals exposed to various forms of trauma.^22-29^ In non-postpartum samples, responses on the PDI predict subsequent PTSD symptoms severity and may be promising in informing subsequent PTSD diagnosis.^16,30,31^ This suggests that responses on the PDI may offer important information in support of screening for CB-PTSD.

Currently, evidence supporting the PDI’s clinical potential to screen for CB-PTSD is lacking. Although several studies reveal that maternal CB-PTSD symptom severity is strongly and positively associated with a woman’s PDI score related to recent childbirth,^1,21^ no study has established a clinical cutoff value to inform the detection and prediction of women with CB-PTSD.

To this end, we studied a sample of 3,039 women who recently gave birth. First, we investigated the factorial structure of the PDI to reveal its underlying constructs. Second, we determined the optimal cutoff value for identifying CB-PTSD in postpartum women.

## Methods

### Sample

This study is part of an investigation of the impact of childbirth experience on maternal mental health.^1,21,32,33^ Women age 18+ years who gave birth to a live baby in the last six months were enrolled and provided information about their mental health and childbirth experience via an anonymous web survey. Recruitment was conducted over the periods of November 2016 to July 2018 and April 2020 to December 2020, using hospital announcements, social media, and professional organizations. The sample in this study consists of 3,039 participants who provided responses on the PDI and PCL-5. The project received exemption from the Partners Healthcare (Massachusetts General Brigham) Human Research Committee (PHRC).

In this sample (n=3,039), mean maternal age was 31.9 years (SD=4.6). The majority were married (92.8%), employed (73.9%), and completed a bachelor’s degree or higher (77.0%). Around half (54.9%) were primiparous, and the majority engaged in skin-to-skin contact (86.9%) and roomed in with their infant (90.9%). 92.5% gave birth full-term, and 70.0 % delivered vaginally. 16.8% underwent emergency or unplanned Caesarean section.

### Measures

The Peritraumatic Distress Inventory (PDI)^22^ is a 13-item self-report questionnaire measuring the degree of emotional and physiological distress endorsed during and shortly after a specified traumatic event. Items are rated on a 5-point Likert scale: 0: Not at All True; 1: Slightly True; 2: Somewhat True; 3: Very True; and 4: Extremely True. The items are summed to achieve total scores in the range 0-52. The PDI shows strong reliability and validity in non-postpartum samples and in the current study (Cronbach’s α = 0.873).

The PTSD Checklist for DSM-5 (PCL-5)^34^ is a 20-item self-report questionnaire measuring the presence of the PTSD DSM-5 symptoms^35^ and their severity. It is the standard measure recommended by the Veterans Affairs (VA) Medical Center for provisional diagnosis of PTSD.^36^ Items are rated on a 5-point Likert scale: 0: Not At All; 1: A Little Bit; 2: Moderately; 3: Quite A Bit; and 4: Extremely. The items are summed to obtained total scores ranging from 0-80. The PCL-5 demonstrates excellent psychometric properties and strong correspondence with diagnostic clinician interview.^36,37^ Reliability is high in the current sample (Cronbach’s α = 0.934). In accord with DSM-5 PTSD classification,^35^ participants were also asked to report the degree to which their symptoms caused impairment in functioning on a 5-point Likert scale. We classified individuals with high probability of CB-PTSD diagnosis, i.e., “positive PTSD screen”, based on (1) the suggested cutoff of 33,^36^ and (2) significant impairment in functioning (i.e., score of 2+ per DSM-5 classification). A negative PTSD screen indicating individuals as having a low probability of CB-PTSD diagnosis was determined if PCL-5<=5 and impairment in functioning < 2.

## Data Analysis

### (i) Exploratory Graph Analysis

To examine the factorial structure of the PDI when used to assess childbirth trauma, we employed Exploratory Graph Analysis (EGA)^38^ using the *EGAnet* R package,^39^ a network psychometrics method that uses undirected network models to assess the psychometric properties of questionnaires. EGA was used to verify the number of factors, and the items associated with each factor, via a graphical lasso.^40^ Network loadings, roughly equivalent to factor loadings, are reported using the *net*.*loads* function, with suggested general effect size guidelines for network loadings of 0.15 for small, 0.25 for moderate, and 0.35 for large.^41^ Next, to examine the stability of the EGA, we followed the analysis with Bootstrap Exploratory Graph Analysis with 5,000 resampling cycles. Finally, we used the *itemStability* function to detect unstable items that hindered the factorial structure stability; unstable items switch factors in more than 25% of the iterations. We re-conducted the EGA without such items. The quality of fit of the final PDI version was estimated using confirmatory factor analysis (CFA) via the *lavaan* Structural Equations Modeling (SEM) package.^42^

### (ii) Utility of the PDI as a screening test for CB-PTSD

Next, we examined the effectiveness of the revised PDI (see Results section) in differentiating between participants with high probability of having CB-PTSD (a score of 33+ for symptom severity on the PCL-5, and a score of 2+ in impairment in functioning) and those with a low probability of having CB-PTSD (PCL-5 <=5, and a score <2 in impairment in functioning). To do so, we calculated the optimal clinical cut-point by bootstrapping the optimal cut-point while maximizing the sensitivity and specificity (i.e., highest Youden’s index:^43^ sensitivity + specificity – 1). We also reported the suggested indexes of the “number needed to diagnose” (NND),^44^ the number of patients who need to be examined to correctly detect one person with the disorder of interest in a study population of persons with and without the known disorder; “number needed to misdiagnose” (NNM),^45^ the number of patients who need to be tested in order for one to be misdiagnosed by the test; and the “likelihood to be diagnosed or misdiagnosed” (LDM),^46^ with higher values of LDM (>1) suggesting that a test is more likely to diagnose than misdiagnose.

## Results

### (i) Exploratory Graph Analysis

The EGA network results are presented in Figure 1, and network loadings are listed in Table 1. Our analysis indicates that the PDI’s factorial structure comprises two factors: “Negative Emotions” (8 items), and “Bodily Arousal and Threat Appraisal” (5 items) (with Item 4, “I felt afraid for my own safety”, showing small-to-moderate cross-loading). When appraising the stability of the EGA by bootstrapping with 5,000 resampling cycles, the analysis indicated moderate-to-high stability: SE = 0.45, with confidence interval (CI) for the number of factors ranging from 1.12 to 2.87. The 2-factor solution was prevalent in 72.5% of the bootstrap samples, with 27.5% producing a 3-factor solution. By examining whether a lack of adequate item stability underlies the less-than-ideal factor solution (i.e., prevalence <75% of the leading solution), we found that Item 4 has poor stability (Figure 2). Hence, we omitted that item and re-conducted the EGA and bootstrap EGA, which revealed perfect factorial and item stability (SE = 0; 100% 2-factor solution). A confirmatory factor analysis (CFA) was used to corroborate the EGA solution, and verify the factorial structure, *CFI* = 0.99, *TLI* = 0.98, *RMSEA* = 0.066 (90% confidence interval [CI] of 0.062, 0.069), *SRMR* = 0.053. The CFA is presented in Figure 3 and shows that the two factors of PDI correlate strongly, *r* = 0.66.

**Figure 1.**
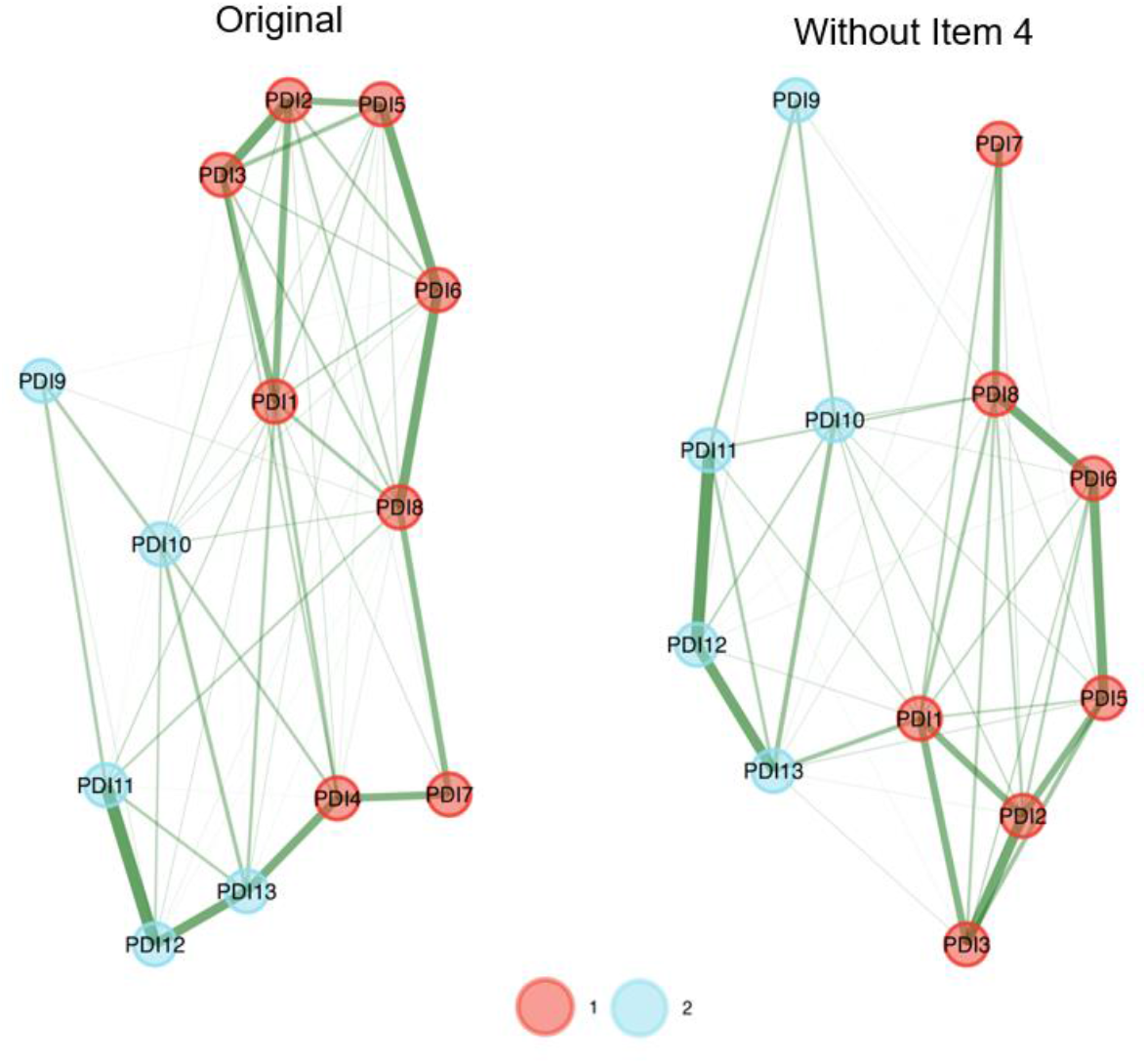
Exploratory Graph Analysis (EGA) results of the original (left network) and the final (right network) Peritraumatic Distress Inventory (PDI) version. The factorial structure of the PDI comprises two factors: Factor 1 (red): Negative Emotions, and Factor 2 (blue): Bodily Arousal and Threat Appraisal. Thicknesses of lines (edges) indicate the strength of association.

**Table 1A:**
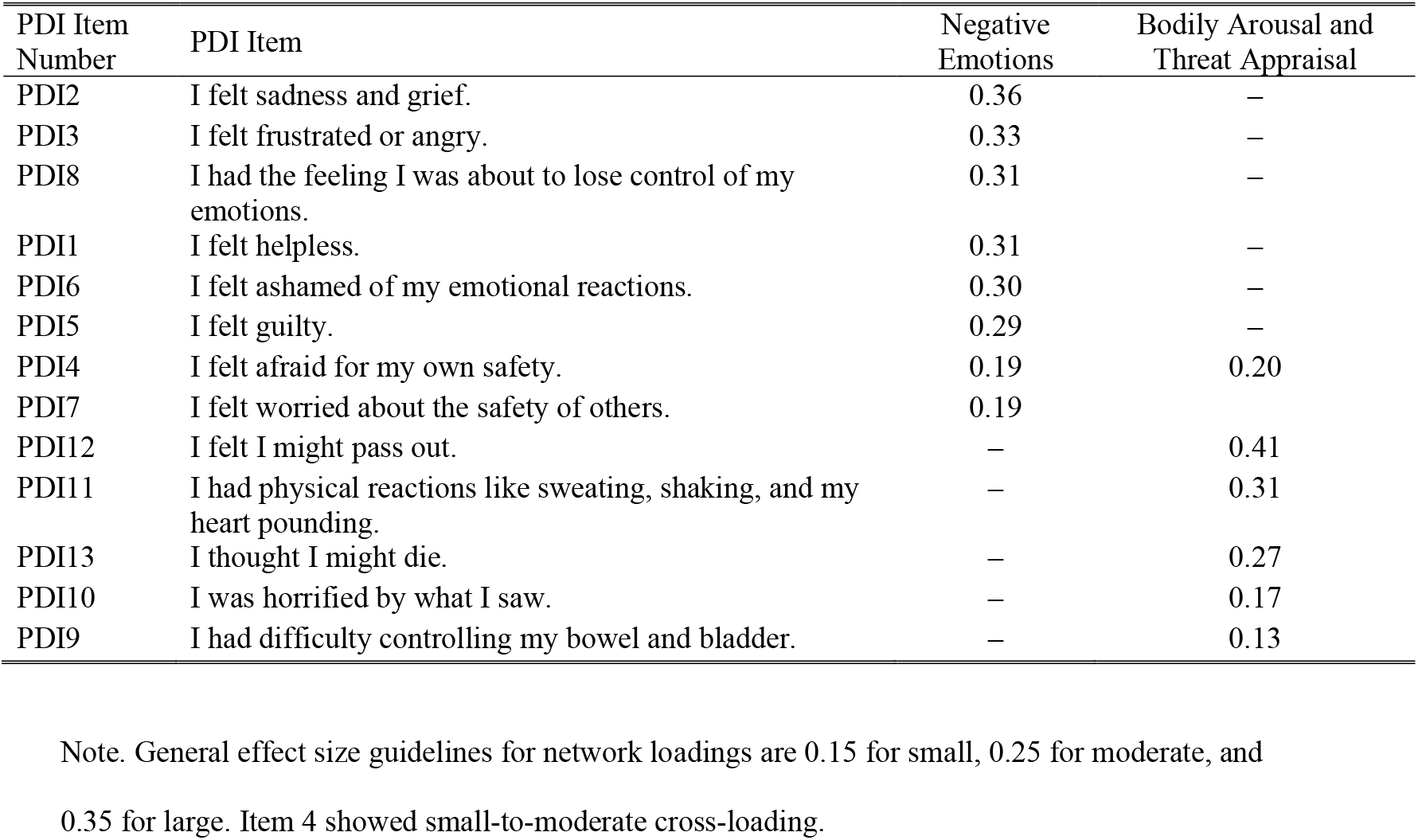
Network loadings based on the Exploratory Graph Analysis (EGA) of the original Peritraumatic Distress Inventory (PDI)

**Table 1B:** Network loadings based on the Exploratory Graph Analysis (EGA) of the revised Peritraumatic Distress Inventory (PDI)

0.35 for large. Item 4 showed small-to-moderate cross-loading.
| PDI Item Number | PDI Item | Negative Emotions | Bodily Arousal and Threat Appraisal |
| --- | --- | --- | --- |
| PDI2 | I felt sadness and grief. | 0.38 | — |
| PDI8 | I had the feeling I was about to lose control of my emotions. | 0.34 | — |
| PDI3 | I felt frustrated or angry. | 0.33 | — |
| PDI6 | I felt ashamed of my emotional reactions. | 0.32 | — |
| PDI1 | I felt helpless. | 0.32 | — |
| PDI5 | I felt guilty. | 0.31 | — |
| PDI7 | I felt worried about the safety of others. | 0.14 | — |
| PDI12 | I felt I might pass out. | — | 0.42 |
| PDI11 | I had physical reactions like sweating, shaking, and my heart pounding. | — | 0.32 |
| PDI13 | I thought I might die. | — | 0.31 |
| PDI10 | I was horrified by what I saw. | — | 0.19 |
| PDI9 | I had difficulty controlling my bowel and bladder. | — | 0.13 |

**Figure 2.**
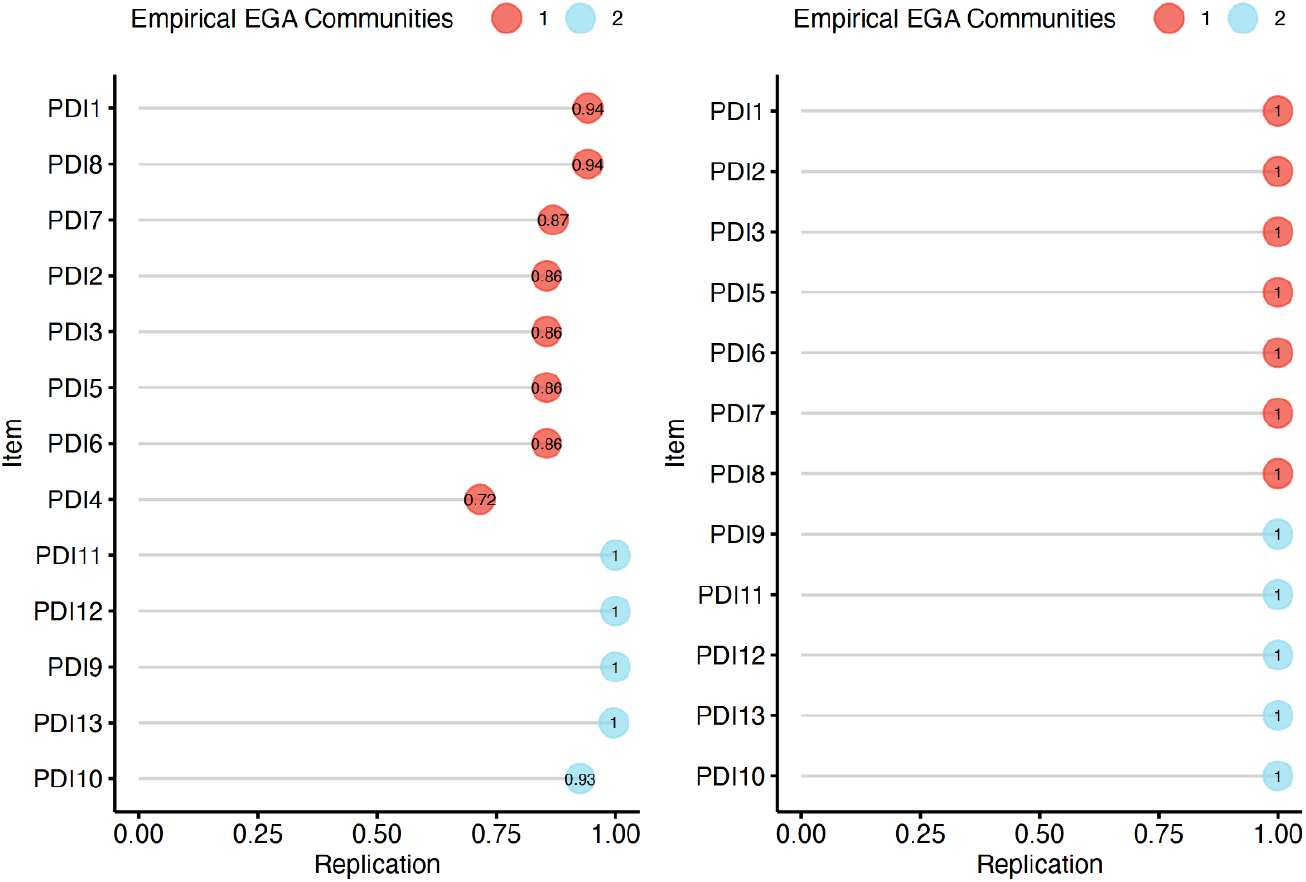
Item stability of the original (left) and revised (right) Peritraumatic Distress Inventory (PDI). Stability below 75% is poor. The factorial structure of the PDI comprises two factors: Factor 1 (red): Negative Emotions, and Factor 2 (blue): Bodily Arousal and Threat Appraisal.

**Figure 3.**
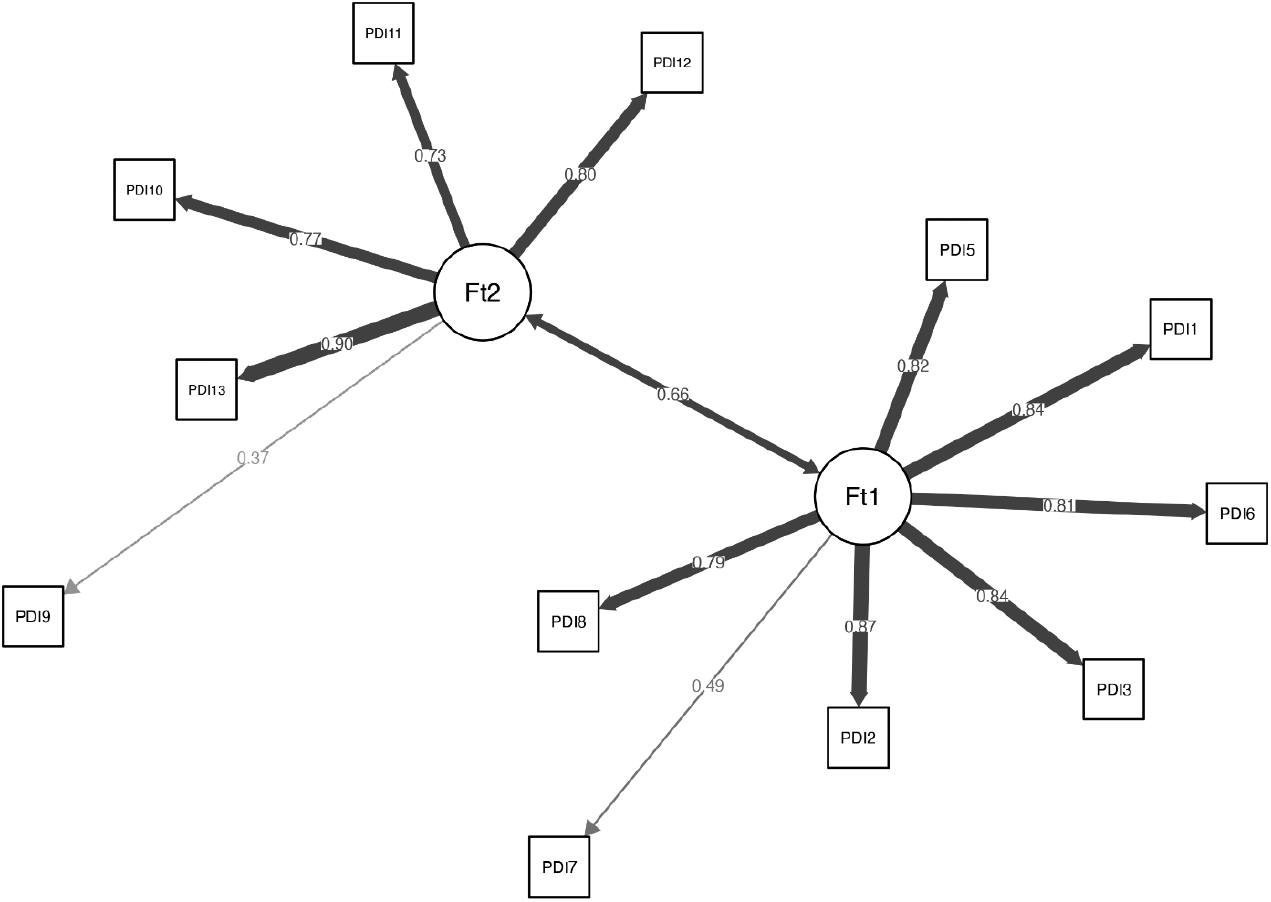
The final confirmatory factor analysis (CFA) for the revised Peritraumatic Distress Inventory (PDI). Ft1 indicates Factor 1: Negative Emotions, and Ft2 indicates Factor 2: Bodily Arousal and Threat Appraisal. Thicknesses of lines (edges) indicate the strength of association.

### (ii) Utility of the PDI as a screening test for CB-PTSD

In this study, 290 participants (9.54%) were classified as having high probability of CB-PTSD (PCL-5 score >=33 and score of >=2 in impairment), and 209 as having low probability of CB-PTSD (PCL-5 <=5 and impairment score <2). Bootstrapping the optimal cut-point of the revised PDI revealed that a cut-point of 15 produces a maximum Youden’s index^43^ of 0.81, with sensitivity of 88.28% and specificity of 92.82% (Figure 4). Using the revised PDI version and cut-point of 15, 1.23 patients are needed to be examined to correctly detect one person with the disorder of interest in a study population of persons with and without the known disorder (i.e., NND value, with 1 as the best possible value). Additionally, 10.18 patients need to be tested for one person to be misdiagnosed by the test (i.e., NNM value). The overall likelihood of being diagnosed compared with being misdiagnosed (i.e., LDM) is 8.26, which indicates high effectiveness in the diagnosis process.

**Figure 4.**
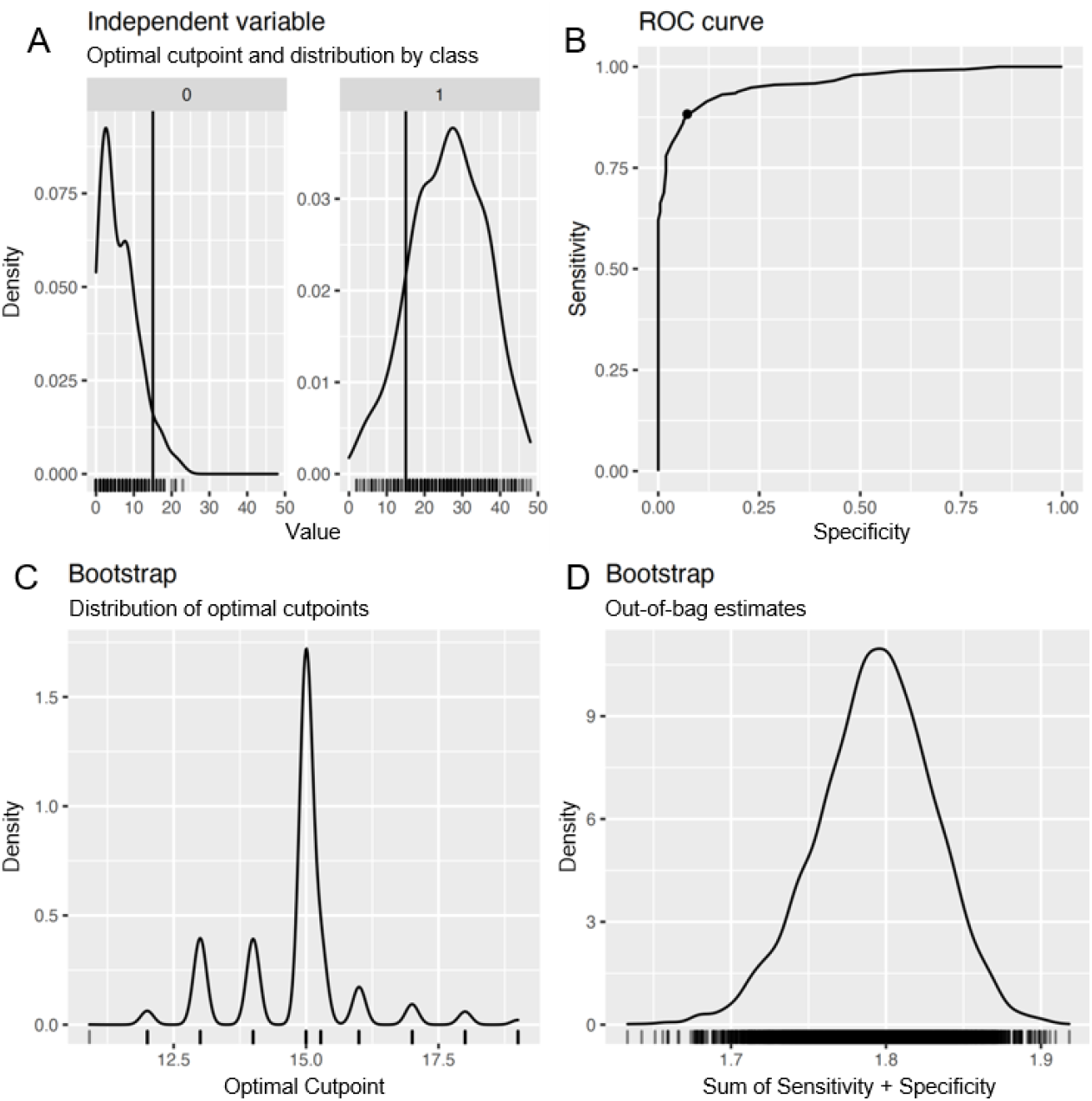
Results of the cut-point optimization process using bootstrap analysis. Panel A: Density of participants below and above the suggested cut-point of 15 in the revised Peritraumatic Distress Inventory (PDI) among the high (1) and low (0) CB-PTSD probability groups. Panel B: The Receiver Operating Characteristic (ROC) curve for the estimation process of the optimal cut-point, with the black dot indicating the highest Youden’s index. Panel C: Density of the optimal cut-point in the estimation process. Panel D: Density of the highest summed sensitivity and specificity scores of the revised PDI.

## Comment

### 1. Principal Findings

This study shows that the Peritraumatic Distress Inventory (PDI) used to assess acute distress in response to recent childbirth can inform identification of women who may endorse PTSD after childbirth (CB-PTSD). A score of 15 on our revised PDI (excluding Item 4, “I felt afraid for my own safety ‘) produced an optimal clinical cutoff associated with high sensitivity and specificity for CB-PTSD endorsement. Our analysis shows that 88% of women who likely meet criteria for CB-PTSD in the first months postpartum will be correctly identified based on their PDI score of 15+. This cutoff results in 8 times greater likelihood of a woman with CB-PTSD being identified rather than being missed based on her PDI screen. Additionally, 93% of women who likely do not meet CB-PTSD criteria will have a score <15.

Our results reveal that the PDI, originally designed to assess emotional and physiological responses experienced during and immediately after a traumatic event, is an effective tool to assess acute distress stemming from the experience of childbirth. Exploratory factor analysis reveals two strongly correlated stable factors consisting of (1) negative emotions and (2) bodily arousal and threat appraisal related to childbirth. Hence, the PDI shows strong construct and convergent validity in postpartum assessment.

### 2. Results

This study is the first investigation of the PDI’s clinical utility to screen for CB-PTSD. Prior studies involving individuals exposed to other forms of trauma suggest that responses on the PDI can identify those likely to endorse PTSD.^25,31,47,48^ Our results advance the literature by demonstrating the PDI’s potential use to screen women for childbirth-related traumatic stress reactions. We establish a clinical cutoff value of 15, designed for the postpartum population, derived from 12 items of the PDI. This cutoff falls in the range of reported cutoffs in non-postpartum samples for the 13-item PDI (cutoffs of 14-23).^25,28,31,47,48^ Our cutoff value suggests that women with CB-PTSD experience childbirth, commonly viewed as a happy event, with elevated distress levels at an magnitude similar to those of individuals who will develop PTSD following other forms of trauma.

Our findings reveal a two-factor structure consistent with the initial validation of the PDI in a sample of police officers who experienced/witnessed various traumatic events^22^ and later studies of trauma-exposed non-postpartum individuals.^25,26^ The underlying factors are largely consistent with previous studies in support of an emotional (distress) component, as well as a cognitive (negative appraisal) factor coupled with hyper-physiological reactivity. The accuracy of our PDI specified to childbirth (sensitivity of 88%) accords with prior studies in non-postpartum samples, with sensitivity rates ranging from 70% to 90%.^25,28,31,47,48^

### 3. Clinical Implications

CB-PTSD is an underrecognized maternal mental health disorder affecting millions of women globally each year.^6^ Early signs of CB-PTSD may go undetected in postpartum women, as recommendations for mental health screening in hospitals and clinics in the U.S. encompass only peripartum depression. Evidence suggests that screening alone for maternal mental health conditions can have benefits,^49^ and treatment may offer maximum benefit and reduce the duration of illness.^50^

Here, we document that the PDI, a brief and simple patient self-report, when used to assess immediate distress reactions to childbirth, can differentiate between individuals with and without CB-PTSD. Accordingly, it may serve as an initial feasible and cost-effective assessment screening before an in-depth diagnostic assessment is performed by a mental health professional.

Because the PDI can be administered in the acute post-trauma period, this screening could potentially be completed during maternity hospitalization stay. This, in turn, may direct high-risk women to receive early interventions or follow-up services, and overcome a major challenge in screening post-discharge, involving the low attendance of follow-up postpartum visits.^51^

Our findings are consistent with research on the role of the subjective elements of trauma in influencing how individuals cope psychologically in the aftermath.^19^ Objective physical morbidity in childbirth and cases of near-miss strongly increase the risk for CB-PTSD;^52^ nevertheless, how the event is experienced and appraised subjectively may profoundly determine maternal mental health outcome.^53,54^ Maternal appraisal could be influenced by perceived support, or lack thereof, during labor and delivery. Hence, the results of our study underscore the importance of promoting positive appraisals and protecting against negative appraisals of childbirth. This suggests opportunities for refining clinical care standards.

### 4. Research Implications

Our study provides proof of concept that the PDI could aid in universal screening of maternal CB-PTSD. We included postpartum women with medically complicated and uncomplicated deliveries, and our recommended clinical cutoff of 15 is derived from this sample. It remains unclear whether a higher cutoff value may distinguish between women who are likely or not to develop CB-PTSD following complicated deliveries. Replication in high-risk groups, e.g., women with severe maternal morbidity (SMM), remains to be done. Although responses on the PDI are consistent in test-retest,^22,24,25^ future research is warranted to determine the PDI’s predictive value when administered soon after childbirth to inform clinical recommendations.

### 5. Strengths and Limitations

This study reveals for the first time the potential clinical utility of the PDI as a screening tool for CB-PTSD. Complementing previous work that has included other, non-childbirth trauma when assessing the PDI’s properties, our study uses this instrument to measure women’s reactions to their recent childbirth, thus informing our understanding of this measure as applied to a postpartum population. Our sample size of 3,039 women is relatively large, enabling conclusions to be drawn with high confidence for our study population.

Several limitations of this study should be noted. We used a cross-sectional study design using a single time point of data collection, and our sample involved a relatively homogeneous population consisting mainly of middle-class Caucasian women in the U.S. Future work is warranted to examine the predictive utility of the PDI administered as closely as possible to the time of childbirth. Clinical cutoffs should be established in ethnically and racially diverse populations. Although we determined the likelihood for CB-PTSD using the well-validated PCL-5 patient self-report, which strongly accords with the clinician-administered PTSD Scale for DSM-5,^55^ we did not perform clinical assessment to confirm diagnosis.

### 6. Conclusions

This study provides evidence that the Peritraumatic Distress Inventory (PDI) is a useful tool to address the critical clinical gap in assessing early signs of psychiatric maternal morbidity associated with traumatic childbirth. We report that women who are likely to endorse CB-PTSD can be accurately identified based on their subjective experience of childbirth as assessed with the PDI. This instrument could be feasible to implement in the clinic as a low-cost, low-burden screening method. This may serve as an initial step in complementing the recommended routine screening for peripartum depression,^49^ in alignment with the recommendation of the American College of Obstetricians and Gynecologists (ACOG) to conduct a full mental health assessment as part of a comprehensive postpartum visit.^56^ As maternal morbidity triggered by life-threatening childbirth-related events poses a significant public health concern in the U.S., our findings may inform early CB-PTSD screening in postpartum maternal populations, to advance progress toward the goals of efficient diagnosis and effective treatment for this disorder.

## Data Availability

All data used in the present study are available upon reasonable request to the authors.

## Acknowledgments

The authors thank Gabriella Dishy (Columbia Psychiatry and New York State Psychiatric Institute) and Rasvitha Nandru (University of Massachusetts Memorial Medical Center) for assisting with data collection. Neither person received funding for this work.

